# Acute and Subacute Ischemia of Lower Limbs: Angiography and Peripheral Hemodynamics

**DOI:** 10.1101/2023.02.21.23285736

**Authors:** Miroslav Bulvas, Zuzana Sommerová

## Abstract

**Objective:** To identify pretreatment characteristics related to ischemia severity and symptoms duration in patients with acute (ALI) and subacute lower limb ischemia (SLI).

**Methods:** Comparative study, a part of prospective, single-arm, single-center trial of 316 consecutive patients with threatened extremities (all-comers, mean age: 70.9±12.0 years, range: 23-96; 184 men) of whom 99. 4 % suffered from acute or subacute supratibial occlusions (target occlusions).

**Results:** Multisegmental (supratibial plus infrapopliteal) arterial occlusions were documented in 90.6 % of ALI patients and in 78.1 % of SLI patients with threatened lower extremity. The value of ABI related to the number of occluded tibial arteries. Occlusion locations associated with ALI (IIA+IIB) were the aortoiliac segment and/or profunda femoris. Furthermore, ALI subjects presented additional infrapopliteal occlusion (acute and/or subacute), smaller number of patent tibial arteries and lower ABI.

**Conclusion:** Anatomically, the most important factor related to the ischemia severity and symptoms duration was the status of infrapopliteal vascular bed. The number of acutely, subacutely or chronically occluded tibial vessels was related to clinical categories of ALI and SLI and to ABI. This suggests that tibial reperfusion can be essential for the salvage of threatened limb and should be always considered.

## INTRODUCTION

Acute lower limb ischemia (ALI) is a critical vascular emergency that endangers the affected extremity and puts the patient’s life at risk. It is defined as a sudden decrease in or worsening of limb perfusion that has been present for 14 days or less and causing a potential threat to limb viability^1,2^.

The clinical presentation of ALI is not single and varies from viable to irreversibly damaged extremity. The limb can be threatened immediately after the symptoms origin or later when ischemia progresses, steadily or in steps. The fact that suddenly developed ischemia can escalate during the time period longer than 2 weeks, led many authors to use the term “subacute limb ischemia” (SLI) to distinguish the unstable and developing condition from the chronic, stable ischemic status. The guidelines^3,4^ defined SLI by symptoms duration longer than 14 days, and lasting up to 3 months.

The pre-treatment factors, reported in the literature^5,6^ to influence the mortality and amputation rates were high age, female gender, non-Caucasian race, comorbidities (diabetes, malignancy, neurological disorders, coronary heart disease, congestive heart disease), multilevel occlusion, more proximal occlusions, limb ischaemia severity, treatment risk and urgency, tissue damage, repeated ALI attacks, absence of early anticoagulation and delay in diagnosis and revascularization. However, their role and incidence may differ in patients with different clinical presentation.

Because the same initial treatment techniques (open surgery, catheter-directed thrombolysis, mechanical thrombectomy) are used in patients with ALI or SLI ^7^, different number of patients with SLI and those with ALI categorized as type I (viable extremity), IIA (marginally threatened) or IIB (immediately threatened) have been often grouped in ALI cohorts in studies to date analyzing surgical and/or endovascular treatment outcomes. To understand whether such a grouping can affect the overall results and comparability among the studies, it is important to learn more about characteristic of individual clinical categories. Closer knowledge is necessary for better understanding the high mortality and amputation rates, and the factors associated with or responsible for clinical presentation, serious events and technical failure of therapeutic intervention.

The aim of this study was to identify the relations of ischemia severity and/or symptoms duration with age, gender, comorbidities, clinical presentation, arterial occlusion aetiology, vascular anatomy and peripheral hemodynamics in patients with threatened lower extremity due to ALI (IIA, IIB) or SLI. Specifically, we focused on the role of infrapopliteal arterial patency in 316 patients (all-comers) of whom 99. 4 % primarily suffered from acute or subacute supratibial occlusions (target occlusions).

## MATERIAL AND METHODS

Our study was a single-centre, prospective, single-arm trial performed during the time interval 2009–2015 with two main goals. The first part documented the efficacy, safety and applicability of endovascular removal of occlusive material as the initial treatment in patients with limb threatening ALI and SLI. It was already published ^8^.

In this report, we focused on pre-treatment difference between clinical categories (ALI vs. SLI, ALI IIB vs. ALI IIA, females vs. males, thrombotic occlusion vs. embolism). We compared demographic data, frequencies of risk factors and comorbidities, clinical data, ankle-brachial pressure index (ABI) and vascular anatomy characteristics (Table 1, 2).

**Table 1.**
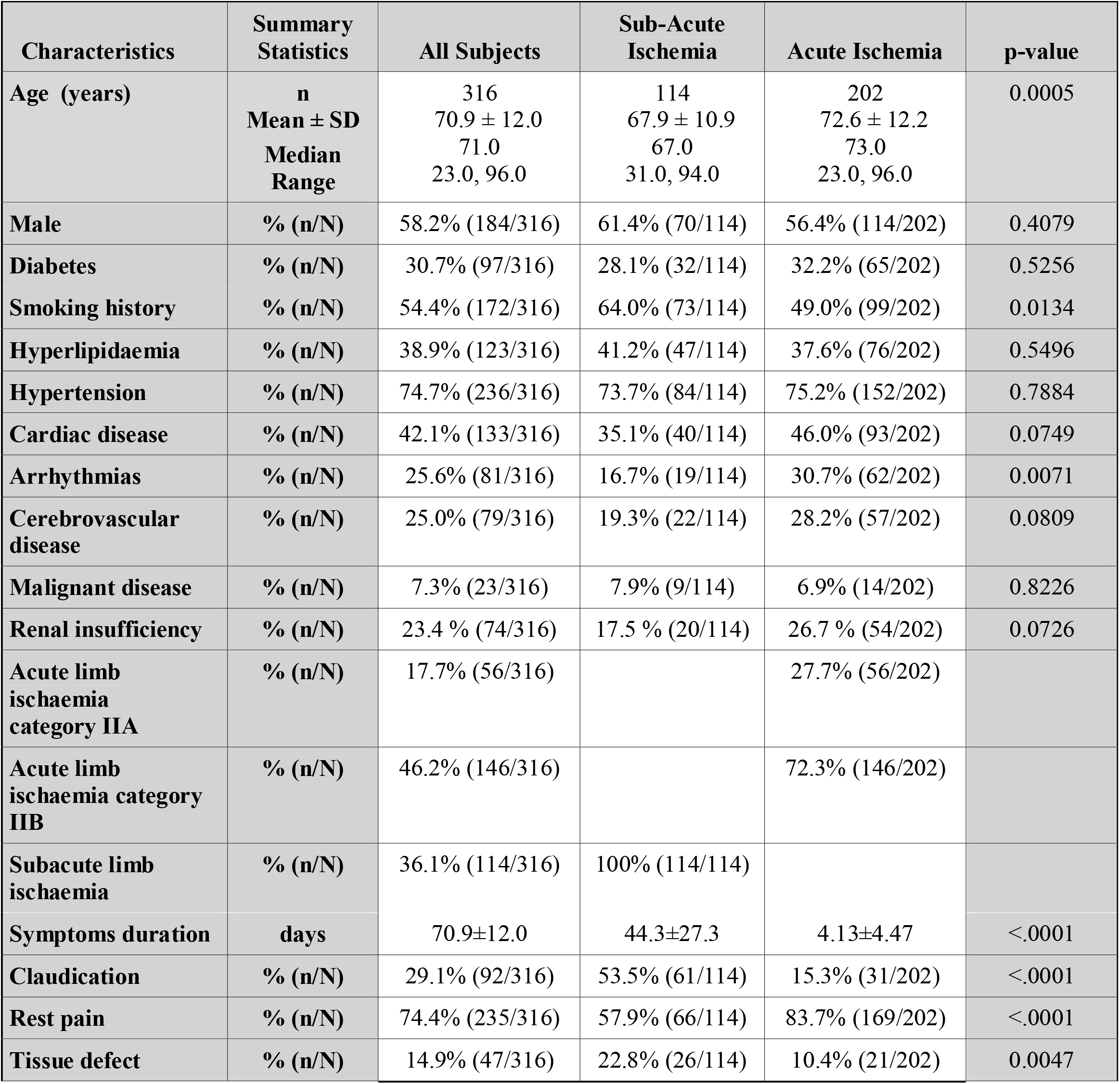
Demography, concomitant disease, ischemia classification and symptoms: SLI vs. ALI

**Table 2.** Hemodynamics, target occlusion cause and location, patent calf vessels: SLI vs. ALI

| Characteristics | Summary Statistics | All Subjects | Sub-Acute Ischemia | Acute Ischemia | p-value |
| --- | --- | --- | --- | --- | --- |
| **Target occlusion length (mm)** | **n**<br>**Mean ± SD**<br>**Median**<br>**Range** | 316<br>229.2 ± 148.1<br>200.0<br>15.0, 650.0 | 114<br>202.0 ± 136.7<br>150.0<br>15.0, 650.0 | 202<br>244.6 ± 152.4<br>200.0<br>20.0, 590.0 | 0.0159 |
| **Target occlusion cause: Embolism** | **% (n/N)** | 19.0% (60/316) | 8.8% (10/114) | 24.8% (50/202) | 0.0005 |
| **Target occlusion cause: Thrombosis** | **% (n/N)** | 81.0% (256/316) | 91.2% (104/114) | 75.2% (152/202) | 0.0005 |
| **Aortoiliac segment occlusion** | **% (n/N)** | 11.1% (35/316) | 6.1% (7/114) | 13.9% (28/202) | 0.0401 |
| **Femoropopliteal segment occlusion** | **% (n/N)** | 73.1% (231/316) | 78.9% (90/114) | 69.8% (141/202) | 0.0868 |
| **Femoropopliteal bypass occlusion** | **% (n/N)** | 22.8% (72/316) | 17.5% (20/114) | 25.7% (52/202) | 0.1241 |
| **Femoro-femoral cross bypass occlusion** | **% (n/N)** | 0.6% (2/316) | 0 | 1.0% (2/202) | 0.5373 |
| **Leg of bifurcation stentgraft occlusion** | **% (n/N)** | 0.3% (1/316) | 0 | 0.5% (1/202) | 1.0000 |
| **Ileofemoral bypass occlusion** | **% (n/N)** | 0.3% (1/316) | 0.9% (1/114) | 0 | 0.3608 |
| **Deep femoral artery occlusion** | **% (n/N)** | 10.1% (32/316) | 5.3% (6/114) | 12.9% (26/202) | 0.0332 |
| **In-stent occlusion** | **% (n/N)** | 23.4% (74/316) | 21.9 % (24/114) | 24.3 % (49/202) | 0.6800 |
| **Additional infrapopliteal occlusion** | **% (n/N)** | 61.7% (195/316) | 50.0% (57/114) | 68.3% (138/202) | 0.0017 |
| **Number of patent calf vessels before treatment** | **n**<br>**Mean ± SD**<br>**Median**<br>**Range** | 316<br>0.9 ± 1.1<br>0.0<br>0.0, 4.0 | 114<br>1.3 ± 1.2<br>1.0<br>0.0, 3.0 | 202<br>0.7 ± 1.0<br>0.0<br>0.0, 4.0 | <.0001 |
| **ABI before treatment** | **n**<br>**Mean ± SD**<br>**Median**<br>**Range** | 309<br>0.1 ± 0.2<br>0.0<br>0.0, 0.6 | 108<br>0.2 ± 0.2<br>0.3<br>0.0, 0.6 | 201<br>0.1 ± 0.1<br>0.0<br>0.0, 0.6 | <.0001 |

**Table 3.** Demographics, concomitant disease, symptoms and target occlusion cause: ALI IIA vs. ALI IIB

| Characteristics | Summary Statistics | All ALI Subjects | ALI IIA | ALI IIB | p-value |
| --- | --- | --- | --- | --- | --- |
| Age (years) | n<br>Mean ± SD<br>Median<br>Range | 202<br>72.6 ± 12.2<br>73.0<br>23.0, 96.0 | 56<br>70.8 ± 12.9<br>73.0<br>23.0, 96.0 | 146<br>73.2 ± 11.9<br>74.0<br>43.0, 96.0 | 0.2113 |
| Male | % (n/N) | 56.4% (114/202) | 62.5% (35/56) | 54.1% (79/146) | 0.4079 |
| Diabetes | % (n/N) | 32.2% (65/202) | 21.4% (12/56) | 36.3% (53/146) | 0.0452 |
| Smoking history | % (n/N) | 49.0% (99/202) | 60.7% (34/56) | 44.5% (65/146) | 0.0425 |
| Hyperlipidaemia | % (n/N) | 37.6% (76/202) | 50.0% (28/56) | 32.9% (48/146) | 0.0344 |
| Hypertension | % (n/N) | 75.2% (152/202) | 71.4% (40/56) | 76.7% (112/146) | 0.4685 |
| Cardiac disease | % (n/N) | 46.0% (93/202) | 41.1% (23/56) | 47.9% (70/146) | 0.4319 |
| Arrhythmias | % (n/N) | 30.7% (62/202) | 28.6% (16/56) | 31.5% (46/146) | 0.7357 |
| Cerebrovascular disease | % (n/N) | 28.2% (57/202) | 17.9% (10/56) | 32.2% (47/146) | 0.0543 |
| Malignant disease | % (n/N) | 6.9% (14/202) | 5.4% (3/56) | 7.5% (11/146) | 0.7615 |
| Renal insufficiency | % (n/N) | 23.4% (74/316) | 17.5% (20/114) | 26.7% (54/202) | 0.0726 |
| Symptoms duration (days) | n<br>Mean ± SD<br>Median<br>Range | 202<br>4.1±4.5<br>2.0<br>0.04, 14.0 | 56<br>6.8±5.3<br>6.5<br>0.08, 14.0 | 146<br>3.1±3.6<br>2.0<br>0.04, 14.0 | 0.0004 |
| Claudication | % (n/N) | 15.3% (31/202) | 39.3% (22/56) | 6.2% (9/146) | <.0001 |
|  | % (n/N) |  |  |  |  |
| Rest pain | % (n/N) | 83.7% (169/202) | 62.5% (35/56) | 91.8% (134/146) | <.0001 |
| Tissue defect | % (n/N) | 10.4% (21/202) | 10.7% (6/56) | 10.3% (15/146) | 1.000 |
| Target occlusion cause: Embolism | % (n/N) | 24.8% (50/202) | 14.3% (8/56) | 28.8% (42/146) | 0.0442 |
| Target occlusion cause: Thrombosis | % (n/N) | 75.2% (152/202) | 85.7% (48/56) | 71.2% (104/146) | 0.0442 |

The target lesion and vascular anatomy information was obtained by digital subtraction angiography in all patients. No therapeutic interventions were performed or tested in this part of the study.

The study included 316 consecutive patients (all-comers, 184 men, mean age: 70.9 ± 12.0 years): 202 subjects with ALI (114 men, mean age: 72.6 ± 12.2, mean symptoms duration: 4.13 ± 4.47 days; 146 patients categorized as IIB of the Rutherford classification for acute limb ischaemia^2^; 56 patients of the category IIA) and 114 patients with SLI (70 men, mean age: 67.9 ± 10.9 years; mean symptoms duration: 44.3 ± 27.3 days). SLI patients were categorized on the basis of the Rutherford classification for chronic limb ischemia ^2^ (38 patients: category 3; 50 patients: category 4; 26 patients: category 5). Critical limb ischemia was diagnosed in 74 patients with SLI according to established guidelines ^1,9,10^.

Target, supratibial occlusions (acute or subacute) were located in aortoiliac arterial segment, femoropopliteal segment, deep femoral artery, bypasses (aortofemoral, femoropopliteal, femorofemoral) and leg of bifurcation stentgraft (Table 2) in 99.4 % of patients. Isolated infrapopliteal occlusions were detected in two patients (0.6 %).

Occlusion cause was determined on the basis of clinical manifestation, angiographic findings^11^, resistance to guidewire penetration and occurrence of underlying lesion after the clot removal. Thrombotic occlusion was diagnosed in 256 (81.0 %), and embolism in 60 (19.0 %) patients.

The University Hospital Královské Vinohrady and the Third Medical Faculty of the Charles University Centralized Institutional Review Board approved the study protocol and informed consent form (identifier: EK/IV-2/2009). The study was conducted in accordance with Declaration of Helsinki, and all patients provided written informed consent. The trial was registered with the International Standard Randomized Controlled Trials Number registry (ISRCTN 154967770). The inclusion criteria allowed enrolment of adult patients with ALI (symptoms duration ≤ 14 days) or SLI (symptoms duration >14 days and ≤ 3 months), severe claudication, ischemic rest pain, sensorimotor deficit and/or tissue defects. Patients presented from a variety of sources – overflow from other urban institutions, remote community hospitals, physician referral, and self-presenting. The exclusion criteria were irreversible ischemia and viable extremity.

Statistical analysis: continuous data are presented as the mean ± standard deviation (SD). Categorical data are given as the number (percentage). Continuous variables were compared using a paired or unpaired t test or Wilcoxon rank sum test, as appropriate. Categorical variables were analyzed using a 2 -sided chi-square or Fischer exact test, as appropriate. For analysis of variance (ANOVA), Welch’
ss test for unequal sample sizes was used. The threshold of statistical significance was p <0.05. Statistical analyses were performed using SAS software (version 9.4; SAS Institute, USA) and STATSOFT software for Welch’s test (Statistica, version 10, USA).

## RESULTS

### Anatomy

The presence of acute or subacute supratibial arterial occlusions was documented in 314 (99.4 %) of 316 patients with threatened lower limbs due to ALI and SLI. Mean length of supratibial occlusion was greater in group with ALI (244.6 mm) vs. SLI (202.0 mm), p=0.0159 and in patients with thrombotic cause of the occlusion (246.6 mm) vs. embolism (155.0 mm, p<.0001).

The most frequently occluded (acutely or subacutely) locations were the femoropopliteal segment (in 73.1 % of 316 patients) and femoropopliteal bypass (22.8%). Significant difference in frequency of femoropopliteal segment and femoropoplital bypass occlusion was not documented between patients with ALI vs. SLI and ALI IIB vs. ALI IIA (Table 1).

The aortoiliac segment occlusion was detected in 11.1 % of all patients, with higher incidence in those with ALI (13.9 %) vs. SLI (6.1 %, p=.0401) (Table 2). The incidence of the deep femoral artery occlusion was higher in patients with ALI vs. SLI (12.9 vs. 5.3 %, p=0.0332), embolism vs. thrombosis (23.3 vs. 7.0 %, p=.0006) and females vs. males (15.2 vs. 6.5%, p=.0142) (Table 2).

Infrapopliteal occlusion (acute or subacute) additional to occlusion located supratibially was documented in 61.7 % of all patients (ALI + SLI) with higher frequency in patients with ALI (68.3 %) vs. SLI (50.0 %, p=.0017), ALI IIB (72.6 %) vs. ALI IIA (57.1 %, p=.0427) and embolism (73.3 %) vs. thrombosis (59.0 %, p=.0403) (Table 2, 4, 6).

**Table 4.** Target occlusion length, occlusion location, patent calf vessels and ABI: ALI IIA vs. ALI IIB

| Characteristics | Summary Statistics | All Subjects | ALI IIA | ALI IIB | p-value |
| --- | --- | --- | --- | --- | --- |
| **Target occlusion length (mm)** | **n**<br>**Mean ± SD**<br>**Median**<br>**Range** | 202<br>244.6 ± 152.4<br>200.0<br>20.0, 590.0 | 56<br>215.5 ± 150.2<br>150.0<br>20.0, 550.0 | 146<br>255.7 ± 152.3<br>220.0 20.0,<br>590.0 | 0.0938 |
| **Aortoiliac segment occlusion** | **% (n/N)** | 13.9 %<br>(28/202) | 14.3 %<br>(8/56) | 13.7 %<br>(20/146) | 1.000 |
| **Femoropopliteal segment occlusion** | **% (n/N)** | 69.8%<br>(141/202) | 69.6%<br>(39/56) | 69.9%<br>(102/146) | 1.000 |
| **Femoropopliteal bypass** | **% (n/N)** | 25.7 %<br>(52/202) | 25.0 %<br>(14/56) | 26.0 %<br>(38/146) | 1.000 |
| **Leg of bifurcation stentgraft occlusion** | **% (n/N)** | 0.5% (1/202) | 1.8% (1/56) | 0.5% (1/202) | 0.2772 |
| **Ileofemoral bypass occlusion** | **% (n/N)** | 25.7% (52/202) | 25.0% (14/56) | 26.0% (38/146) | 1.000 |
| **Deep femoral artery occlusion** | **% (n/N)** | 12.9% (26/202) | 5.4% (3/56) | 15.8% (23/146) | 0.0597 |
| **In-stent occlusion** | **% (n/N)** | 24.3% (49/202) | 33.9% (19/56) | 20.5% (30/146) | 0.0657 |
| **Additional infrapopliteal occlusion** | **% (n/N)** | 68.3% (138/202) | 57.1% (32/56) | 72.6% (106/146) | 0.0427 |
| **No. of patent calf vessels before treatment** | **n**<br>**Mean ± std**<br>**Median**<br>**Range** | 202<br>0.7± 1.0<br>0.0<br>0.0, 4.0 | 56<br>1.4 ± 1.1<br>1.0<br>0.0, 3.0 | 146<br>0.4± 0.9<br>0.0<br>0.0, 4.0 | <.0001 |
| **ABI before treatment** | **n**<br>**Mean ± SD**<br>**Median**<br>**Range** | 201<br>0.1 ± 0.1<br>0.0<br>0.0, 0.6 | 55<br>0.2 ± 0.2<br>0.2<br>0.0, 0.6 | 146<br>0.0 ± 0.1<br>0.0<br>0.0, 0.4 | <.0001 |

**Table 5.** Demographics, concomitant disease, ischemia classification and symptoms: thrombosis vs. embolism

| Characteristics | Summary Statistics | All Subjects | Thrombosis | Embolism | p-value |
| --- | --- | --- | --- | --- | --- |
| Age (years) | n<br>Mean ± SD<br>Median<br>Range | 316<br>70.9 ± 12.0<br>71.0<br>23.0, 96.0 | 256<br>70.2 ± 11.3<br>70.0<br>43.0, 96.0 | 60<br>73.7 ± 14.2<br>76.0<br>23.0, 94.0 | 0.0054 |
| Male | % (n/N) | 58.2% (184/316) | 63.7% (163/256) | 35.0% (21/60) | <.0001 |
| Female | % (n/N) | 41.8% (132/316) | 36.3% (93/256) | 65.0% (39/60) | <.0001 |
| Diabetes | % (n/N) | 30.7% (97/316) | 30.1% (77/256) | 33.3% (20/60) | 0.6425 |
| Smoking history | % (n/N) | 54.4% (172/316) | 59.8% (153/256) | 31.7% (19/60) | <.0001 |
| Hyperlipidaemia | % (n/N) | 38.9% (123/316) | 39.5% (101/256) | 36.7% (22/60) | 0.7692 |
| Hypertension | % (n/N) | 74.7% (236/316) | 74.6% (191/256) | 75.0% (45/60) | 1.0000 |
| Cardiac disease | % (n/N) | 42.1% (133/316) | 37.5% (96/256) | 61.7% (37/60) | 0.0008 |
| Arrhythmias | % (n/N) | 25.6% (81/316) | 19.5% (50/256) | 51.7% (31/60) | <.0001 |
| Cerebrovascular disease | % (n/N) | 25.0% (79/316) | 26.6% (68/256) | 18.3% (11/60) | 0.2458 |
| Malignant disease | % (n/N) | 7.3% (23/316) | 7.8% (20/256) | 5.0% (3/60) | 0.5874 |
| Renal insufficiency | % (n/N) | 23.4% (74/316) | 23.8% (61/256) | 21.7% (13/60) | 0.8657 |
| Subacute ischaemia | % (n/N) | 36.1% (114/316) | 40.6% (104/256) | 16.7% (10/60) | 0.0005 |
| Acute ischaemia | % (n/N) | 63.9% (202/316) | 59.4% (152/256) | 83.3% (50/60) | 0.0005 |
| Acute ischaemia class IIA | % (n/N) | 27.7% (56/202) | 31.6% (48/152) | 16.0% (8/50) | 0.0442 |
| Acute ischaemia class IIB | % (n/N) | 72.3% (146/202) | 68.4% (104/152) | 84.0% (42/50) | 0.0442 |
| Claudication | % (n/N) | 29.1% (92/316) | 32.4% (83/256) | 15.0% (9/60) | 0.0071 |
| Critical limb ischemia | % (n/N) | 23.4% (74/316) | 26.6% (68/256) | 10.0% (6/60) | 0.0062 |
| Rest pain | % (n/N) | 74.4% (235/316) | 73.8% (189/256) | 76.7% (46/60) | 0.7436 |
| Tissue defect | % (n/N) | 14.9% (47/316) | 17.2% (44/256) | 5.0% (3/60) | 0.0151 |

**Table 6.** Target occlusion length and location, ABI and patent calf vessels: thrombosis vs. embolism

| Characteristics | Summary Statistics | All Subjects | Thrombosis | Embolism | p-value |
| --- | --- | --- | --- | --- | --- |
| Target lesion length (mm) | n<br>Mean ± SD<br>Median<br>Range | 316<br>229.2 ± 148.1<br>200.0<br>15.0, 650.0 | 256<br>246.6 ± 150.4<br>220.0<br>15.0, 650.0 | 60<br>155.0 ± 111.7<br>125.0<br>20.0, 480.0 | <.0001 |
| Aortoiliac segment | % (n/N) | 11.1% (35/316) | 10.2% (26/256) | 15.0% (9/60) | 0.3588 |
| Femoropopl segment | % (n/N) | 73.1% (231/316) | 68.0% (174/256) | 95.0% (57/60) | <.0001 |
| Femoropopl bypass | % (n/N) | 22.8% (72/316) | 27.7% (71/256) | 1.7% (1/60) | <.0001 |
| Femo-femoral cross bypass | % (n/N) | 0.6% (2/316) | 0.8% (2/256) |  |  |
| Leg of bifurcation stentgraft | % (n/N) | 0.3% (1/316) | 0.4% (1/256) |  |  |
| Ileofemoral bypass | % (n/N) | 0.3% (1/316) | 0.4% (1/256) |  |  |
| Deep femoral artery | % (n/N) | 10.1% (32/316) | 7.0% (18/256) | 23.3% (14/60) | 0.0006 |
| In-stent occlusion | % (n/N) | 23.4% (74/316) | 27.7% (71/256) | 5.0% (3/60) | <.0001 |
| Additional infrapopliteal occlusion | % (n/N) | 61.7% (195/316) | 59.0% (151/256) | 73.3% (44/60) | 0.0403 |
| No. of patent calf vessles before treatment | n<br>Mean ± SD<br>Median<br>Range | 316<br>0.9 ± 1.1<br>0.0<br>0.0, 4.0 | 256<br>1.0 ± 1.1<br>1.0<br>0.0, 3.0 | 60<br>0.4 ± 0.9<br>0.0<br>0.0, 4.0 | <.0001 |
| ABI before treatment | n<br>Mean ± SD<br>Median<br>Range | 309<br>0.1 ± 0.2<br>0.0<br>0.0, 0.6 | 250<br>0.14 ± 0.17<br>0.0<br>0.0, 0.6 | 59<br>0.07 ± 0.14<br>0.0<br>0.0, 0.5 | 0.0016 |

Infrapopliteal occlusion (acute, subacute and chronic; segmental or total) in one, two or three tibial vessels was detected in 86.1 % of 316 patients with significant difference between ALI (90.6 %) vs. SLI (78.1 %, p=.0036) subjects. All tibial vessels occluded were documented in 33.3 % of SLI and 62.4 % of ALI patients (p=.0001) (Fig. 1).

**Fig. 1.**
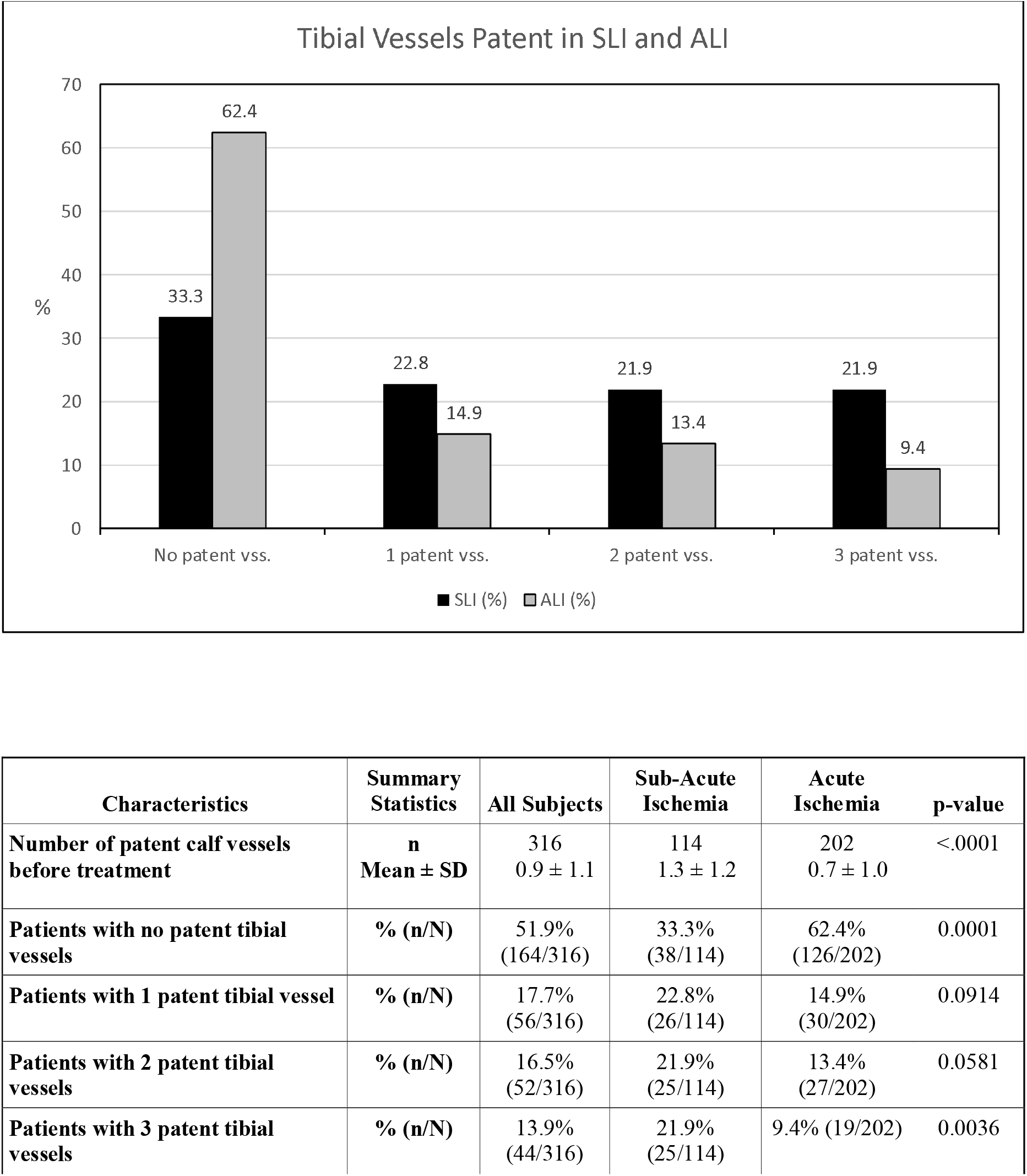
ALI dominates in patients with occlusions (complete or segmental) of all tibial vessels, while SLI majority is present in those with 1, 2 or 3 tibial vessels patent. SD - standard deviation, vss-vessel(s)

Higher mean number of patent (without acute, subacute or chronic occlusions) tibial vessels was detected in patients with SLI (1.3) vs. ALI (0.7, p<.0001), ALI IIA (1.4) vs. ALI IIB (0.4, p<.0001), thrombosis (1.0) vs. embolism (0.4, p<.0001) (Table 2, 4, 6).

Higher incidence of the femoropopliteal segment occlusion was established in subjects with embolism (95.0 %) vs. thrombosis (68.0 %, p<.0001) and in female group (79.5 %) vs. males (68.5 %, p=.0296). Higher frequency of the femoropopliteal bypass occlusion was detected in the male group (28.8 %) vs. females (14.4 %, p=.0027) and patients with thrombotic occlusion (27.7 %) vs. embolism (1.7 %, p<.0001) (Table 6, 7).

**Table 7.** Age, concomitant disease, symptoms, ischemia classification, occlusion length and location: female vs. male gender

| Characteristics | Summary Statistics | All Subjects | Female | Male | p-value |
| --- | --- | --- | --- | --- | --- |
| Age (years) | n<br>Mean ± SD<br>Median<br>Range | 316<br>70.9 ± 12.0<br>71.0<br>23.0, 96.0 | 132<br>75.1 ± 11.4<br>76.5<br>23.0, 96.0 | 184<br>67.9 ± 11.5<br>67.5<br>31.0, 94.0 | <.0001 |
| Diabetes | % (n/N) | 30.7% (97/316) | 33.3% (44/132) | 28.8% (53/184) | 0.3903 |
| Smoking history | % (n/N) | 54.4% (172/316) | 36.4% (48/132) | 67.4% (124/184) | <.0001 |
| Hyperlipidaemia | % (n/N) | 38.9% (123/316) | 41.7% (55/132) | 37.0% (68/184) | 0.4147 |
| Hypertension | % (n/N) | 74.7% (236/316) | 81.1% (107/132) | 70.1% (129/184) | 0.0355 |
| Cardiac disease | % (n/N) | 42.1% (133/316) | 42.4% (56/132) | 41.8% (77/184) | 1.0000 |
| Arrhythmias | % (n/N) | 25.6% (81/316) | 34.8% (46/132) | 19.0% (35/184) | 0.0017 |
| Cerebrovascular disease | % (n/N) | 25.0% (79/316) | 25.0% (33/132) | 25.0% (46/184) | 1.0000 |
| Malignant disease | % (n/N) | 7.3% (23/316) | 6.8% (9/132) | 7.6% (14/184) | 0.8301 |
| Renal insufficiency | % (n/N) | 23.4% (74/316) | 17.4% (23/132) | 27.7% (51/184) | 0.0429 |
| Acute limb ischaemia category IIA | % (n/N) | 27.7% (56/202) | 23.9% (21/88) | 30.7% (35/114) | 0.3420 |
| Acute limb ischaemia category IIB | % (n/N) | 72.3% (146/202) | 76.1% (67/88) | 69.3% (79/114) | 0.3420 |
| Claudication | % (n/N) | 29.1% (92/316) | 23.5% (31/132) | 33.2% (61/184) | 0.0785 |
| Critical limb ischaemia | % (n/N) | 23.4% (74/316) | 21.2% (28/132) | 25.0% (46/184) | 0.5011 |
| Rest pain | % (n/N) | 74.4% (235/316) | 78.8% (104/132) | 71.2% (131/184) | 0.1509 |
| Tissue defect | % (n/N) | 14.9% (47/316) | 9.8% (13/132) | 18.5% (34/184) | 0.0375 |
| Target occlusion cause: Embolism | % (n/N) | 19.0% (60/316) | 29.5% (39/132) | 11.4% (21/184) | <.0001 |
| Target occlusion cause: Thrombosis | % (n/N) | 81.0% (256/316) | 70.5% (93/132) | 88.6% (163/184) | <.0001 |
| Target occlusion length (mm) | n<br>Mean ± SD<br>Median<br>Range | 316<br>229.2 ± 148.1<br>200.0<br>15.0, 650.0 | 132<br>221.0 ± 142.1<br>200.0<br>20.0, 590.0 | 184<br>235.1 ± 152.5<br>200.0<br>15.0, 650.0 | 0.5290 |
| Aortoiliac segment | % (n/N) | 11.1% (35/316) | 14.4% (19/132) | 8.7% (16/184) | 0.1453 |
| Femoropopliteal segment | % (n/N) | 73.1% (231/316) | 79.5% (105/132) | 68.5% (126/184) | 0.0296 |
| Femoropopliteal bypass | % (n/N) | 22.8% (72/316) | 14.4% (19/132) | 28.8% (53/184) | 0.0027 |
| Femo-femoral cross bypass | % (n/N) | 0.6% (2/316) | 0.8% (1/132) | 0.5% (1/184) | 1.0000 |
| Leg of bifurcation stentgraft | % (n/N) | 0.3% (1/316) |  | 0.5% (1/184) | 1.0000 |
| Ileofemoral bypass | % (n/N) | 0.3% (1/316) |  | 0.5% (1/184) | 1.0000 |
| Deep femoral artery | % (n/N) | 10.1% (32/316) | 15.2% (20/132) | 6.5% (12/184) | 0.0142 |
| In-stent occlusion | % (n/N) | 23.4% (74/316) | 24.2% (32/132) | 22.8% (42/184) | 0.7888 |
| Additional infrapopliteal occlusion | % (n/N) | 61.7% (195/316) | 62.9% (83/132) | 60.9% (112/184) | 0.7266 |
| Number of patent calf vessels | n<br>Mean ± SD<br>Median<br>Range | 316<br>0.9 ± 1.1<br>0.0<br>0.0, 3.0 | 132<br>0.9 ± 1.1<br>0.0<br>0.0, 3.0 | 184<br>1.0 ± 1.1<br>0.0<br>0.0, 3.0 | 0.6844 |
| ABI before treatment | n<br>Mean ± SD<br>Median<br>Range | 309<br>0.1 ± 0.2<br>0.0<br>0.0, 0.6 | 129<br>0.12 ± 0.16<br>0.0<br>0.0, 0.6 | 180<br>0.14 ± 0.17<br>0.0<br>0.0, 0.6 | 0.2715 |

### Ankle-brachial index

Generally, the mean ABI was low (0.1) in the group of patients with ALI and SLI before treatment. Its value was higher in patients with SLI (0.22) vs. those with ALI (0.08, p<.0001), in subjects with ALI IIA (0.2) vs. ALI IIB (0.0, p<.0001) and in patients with thrombosis (0.14) vs. embolism (0.07, p=.0016) (Table 2, 4, 6). In subgroup with ABI ranged from 0.0 to 0.1, the patients with ALI were in majority (71.2 %). Patients with SLI dominated in the subgroups with ABI value from 0.2-0.5 (Fig 2). The value of ABI significantly correlated (p<.001) with the number of patent calf vessels in subjects with ALI or SLI (Fig. 3,4).

**Fig. 2.**
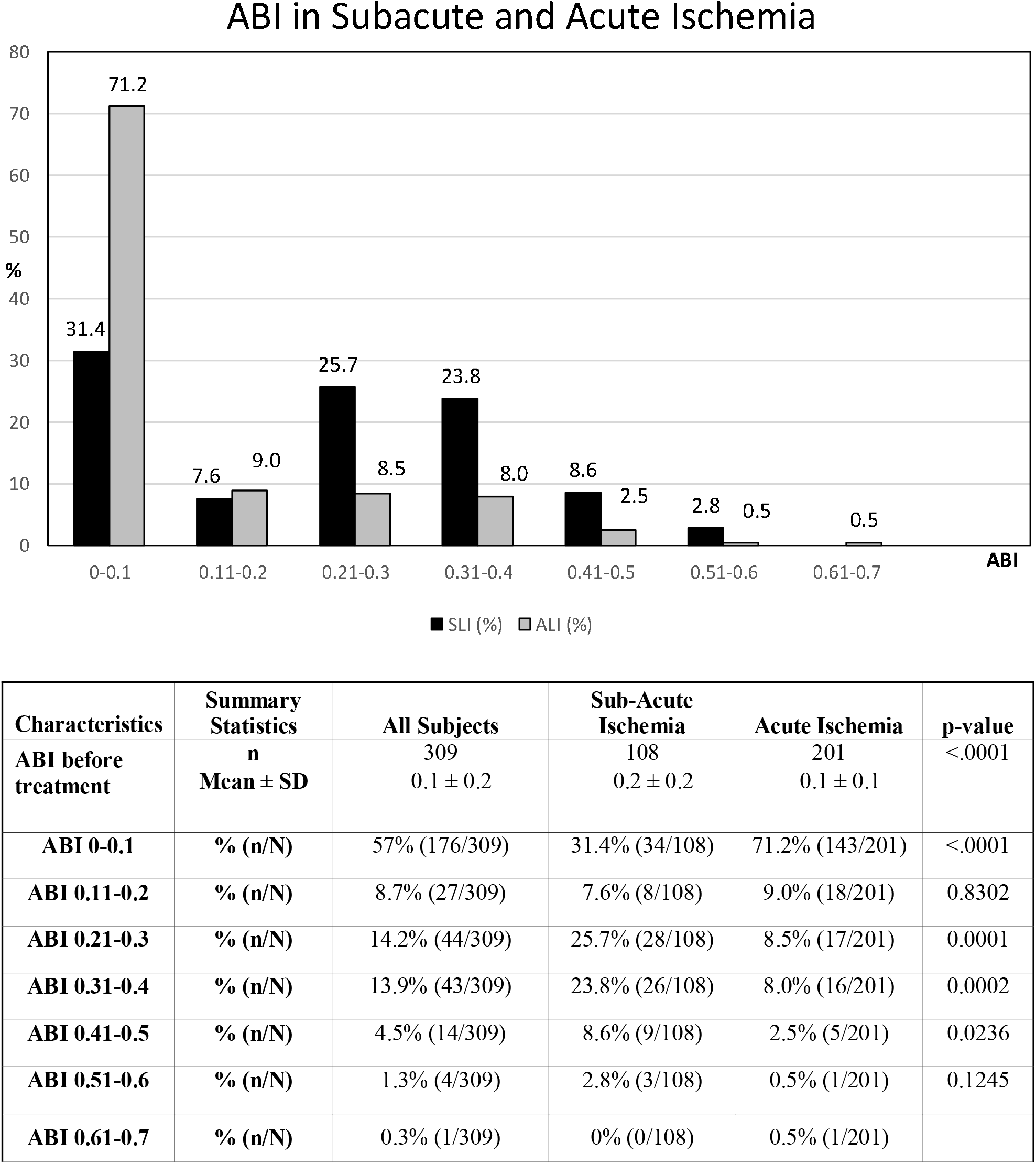
Frequency of SLI and ALI in different ABI intervals. ALI predominates in patients with the lowest ABI (range 0-0.1). In subjects with higher ABI (range 0.21 – 0.5), SLI is more frequent.

**Fig. 3.**
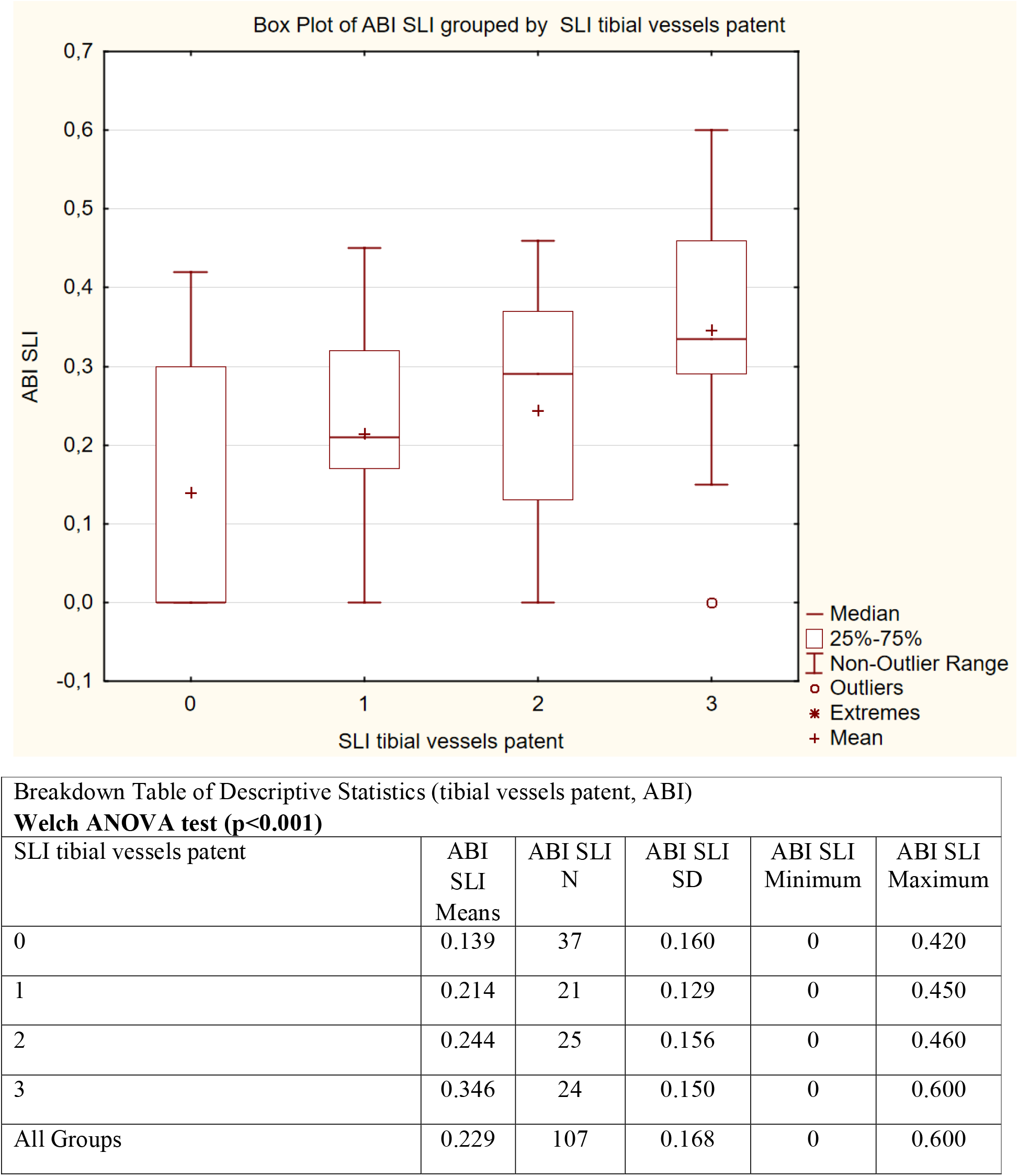
The number of patent tibial vessels significantly (p<.001) correlates to ABI in SLI patients.

**Fig. 4.**
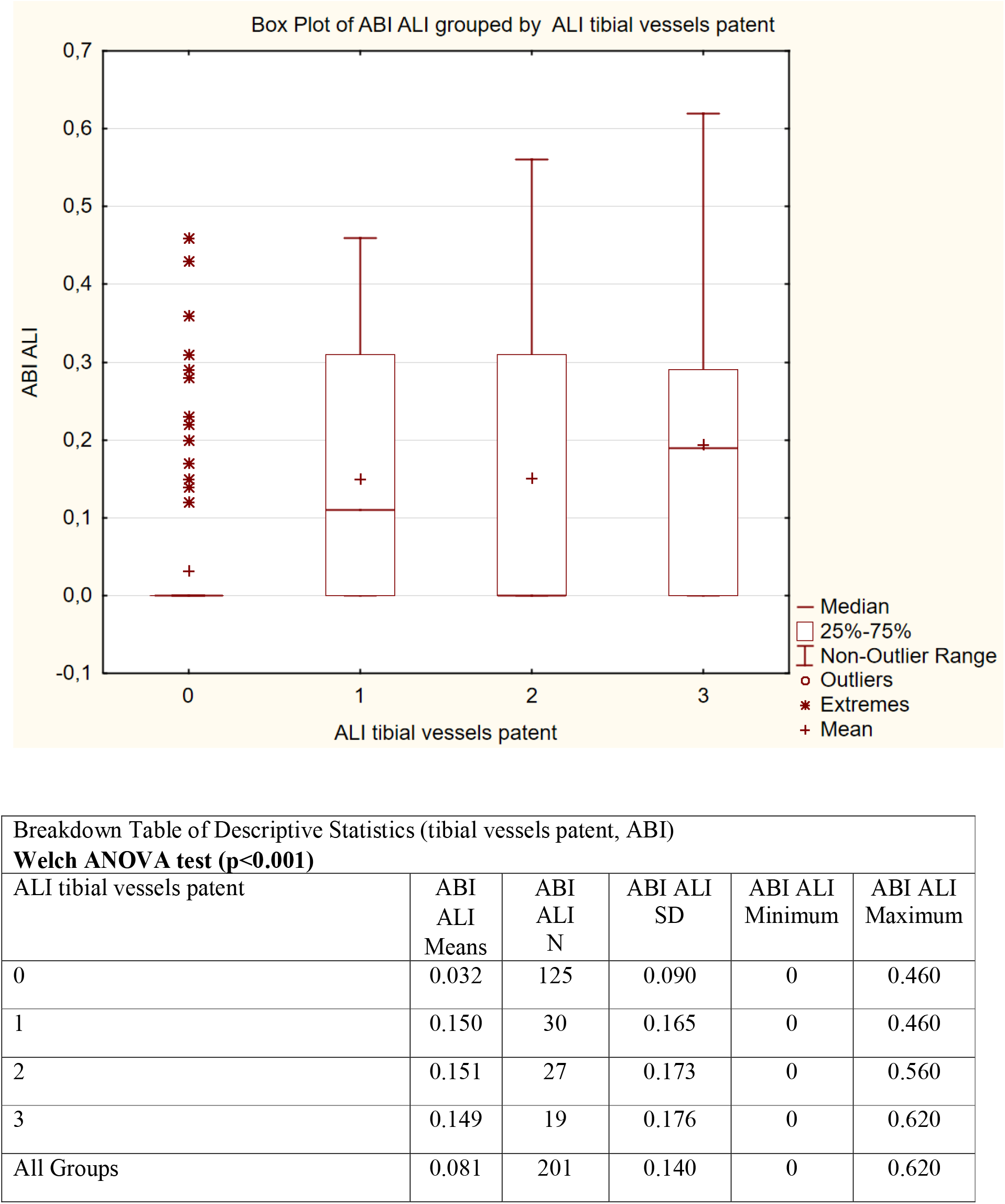
Also in ALI patients, the number of patent tibial vessels significantly (p<.001) correlates to ABI.

### Target occlusion origin: embolism vs. thrombosis

Embolism as the cause of target supratibial occlusion was detected in 19 % of patients with ALI+SLI. This cause was more frequent in patients with ALI (24.8 %) vs. SLI ((8.8 %, p=.0005), ALI IIB (28.8%) vs. ALI IIA (14.3 %, p=.0442) and in females (29.5 %) vs. males (11.4 %, p<.0001) (Table 2, 3, 7).

In comparison with thrombotic lesions, the embolic occlusions were shorter in average (155 vs. 246.6 mm, p<.0001) and they were localized more frequently in the femoropopliteal segment (95.0 vs. 68.0 %, p<.0001) and/or the deep femoral artery (23.3 vs. 7.0 %, p=.0006) (Table 6).

In patients with embolic supratibial closure, higher number (73.3 vs. 59.0 %, p=.0403) of those with additional occlusions (acute, subacute, chronic; embolic or thrombotic) located infrapopliteally was detected. Additionally, these subjects presented lower number of patent calf vessels (0.4 vs. 1.0, p<.0001) and lower ABI (0.07 vs. 0.14, p=.0016).

Embolism was also associated with higher age (73.7 vs. 70.2 years, p=.0054), higher incidence of female gender (65.0 % vs. 36.3 %, p<.0001), cardiac disease (61.7 vs. 37.5 %, p=.0008), arrhythmias (51.7 vs. 19.5 %, p<.0001), ALI (83.3 vs. 59.4 %, p=.0005) and ALI IIB (84.0 vs. 68.4 %, p=.0442).

On the other hand, patients with thrombotic occlusion presented more frequent male gender (63.7 vs. 35 %, p<.0001), positive smoking history (59.8 vs. 31.7 %, p<.0001), SLI (40.6 vs. 16.7 %, p=.0005), ALI IIA (31.6 vs. 16 %, p=.0442), claudication (32.4 vs. 15 %, p=.0071), critical limb ischaemia (26.6 vs.10 %, p=.0062), tissue defect (17.2 vs. 5 %, p=.0151), in-stent occlusion (27.7 vs. 5 %, p<.0001), femoropopliteal bypass occlusion (27.1 vs. 1.7 %, p<.0001) higher number of patent calf vessels (1.0 vs. 0.4, p<.0001) and higher ABI (0.14 vs. 0.07, p=.0016) (Table 5,6).

### Risk factors and concomitant disease

Comparison between patients with SLI and ALI detected significantly higher frequency of the smoking history (64.0 vs. 49.0 %, p=.0134), claudication (53.5 vs. 15.3 %, p<.0001) and tissue defect (22.8 vs.10.4 %, p=.0047). The ALI patients were older (mean age: 72.6 vs. 67.9 years, p=.0005), with longer target lesion (mean length: 244.6 vs. 202.0 mm, p=.0159), higher frequency of rest pain (83.7 vs. 57.9 %, p<.0001), embolism (24.8 vs. 8.8 %, p=.0005), arrhythmias (30.7 vs. 16.7 %, p=.0071). A higher statistical trend toward increased incidence of cardiac disease (46.0 vs. 35.1 %, p=.0749), cerebrovascular disease (28.2 vs.19.3 %, p=.0809) and renal insufficiency (26.7 vs. 17.5 %, p=.0726) was detected in ALI subjects (Table 1,2).

Comparison between ALI patients with immediately (IIB) and marginally (IIA) threatened extremities detected significantly higher frequency of diabetes (36.3 vs. 21.4 %, p=.0452), rest pain (91.8 vs. 62.5 %, p<.0001), embolism (28.8 vs. 14.3 %, p=.0442) in II B subjects, together with lower mean number of patent calf vessels (0.4±0.9 vs. 1.4±1.1, p<.0001), ankle-brachial index (0.0±0.1 vs. 0.2±0.2, p<.0001) and shorter mean duration of the ischaemic symptoms (3.1 vs. 6.8 days, p=.0004). Patients with ALI IIB also presented higher incidence of the cerebrovascular disease (32.2 vs. 17.9 %, p=.0543) and the renal insufficiency (26.7 vs. 17.5 %, p=.0726), but those differences were beyond the statistical significance (Table 3,4). Subjects with ALI IIA presented higher frequency of smoking history (60.7 vs. 44.5 %, p=.0425), hyperlipidaemia (50.0 vs. 32.9 %, p=.0344), claudication (39.3 vs. 6.2 %, p<.0001) and thrombosis (85.7 vs. 71.2 %, p=.0442+) as the cause of target occlusion (Table 3, 4).

### Gender: Females vs. males

Female gender was associated with higher mean age (75.1 vs. 67.9 years, p<.0001), higher incidence of hypertension (81.1 vs. 70.1 %, p=.0355), arrhythmias (34.8 vs. 19.0 %, p=.0017) and embolism (29.5 vs. 11.4 %, p<.0001) as the cause of target arterial occlusion. Additionally, the occlusion was more frequently located in the femoropopliteal arterial segment (79.5 vs. 68.5 %, p=.0296) and the deep femoral artery (15.2 vs. 6.5 %, p=.0142) in this subgroup. Male subgroup presented more frequent smoking history (67.4 vs. 36.4 %, p<.0001), renal insufficiency (27.7 vs. 17.4 %, p=.0429), tissue defect (18.5 vs. 9.8 %, p=.0375), thrombotic cause of the occlusion (88.6 vs. 70.5 %, p<.0001) and femoropopliteal bypass occlusion (28.8 vs. 14.4 %, p=.0027). A higher statistical trend in favour of claudication (33.2 vs. 23.5 %, p=.0785) was also detected in this subgroup (Table 7).

### Clinical presentation

Comparison between ALI patients with immediately (IIB) and marginally (IIA) threatened extremities detected significantly shorter mean duration of the ischaemic symptoms (3.1 vs. 6.8 days, p=.0004). The incidence of tissue defects was higher in males (18.5 % vs. 9.8 %, p=.0375), SLI vs. ALI (22.8 .vs 10.4 %, p=.0047), patients with thrombosis vs. embolism (17. 2 vs. 5.0 %, p=.0151). The rest pain occurred more frequently in ALI group (83.7 % vs. 57.9 %, p<.0001), and in ALI IIB vs. ALI IIA (91.8 vs. 62.5 %, p<.0001). The claudication was detected in 53.5 % of subjects with SLI (vs. 15.3 % in ALI, p<.0001), 39.3 % of ALI IIA (vs. 6.2 % in ALI IIB, p<.0001) and in patients with thrombosis (32.4 %) vs. embolism (15.0 %, p=.0071) (Table 1,3,5,7).

## DISCUSSION

Ischaemia severity, symptoms duration and their progression in time vary with the grade and dynamics of blood flow deterioration. Vessel anatomy before ALI attack, site and cause of acute arterial occlusion, collateral development, embolus fragmentation and its displacement to the peripheral branches, thrombosis progression, runoff quality and antithrombotic therapy are the factors influencing clinical presentation of ALI. Together with serious comorbidities, they affect periprocedural, early and long-term treatment outcomes, morbidity, mortality and amputation rates. Detailed angiography description and pre-treatment ABI data were often not presented in the study reports dedicated to surgical and/or endovascular therapy of ALI. Thus, the correlation of ischaemic symptoms and their duration with the limb hemodynamics and vascular anatomy is difficult or impossible from the literature.

In retrospective analysis of 160 ALI patients (symptoms duration 7 days or less, 13 % embolic and 87 % thrombotic occlusions, 75 % clinical category II) managed with 186 intraarterial thrombolytic procedures, McNamara and colleagues^12^ correlated angiographic patterns with clinical severity of ischemia. They based the angiographic categorization on the number of occluded arterial segments (common iliac, external iliac, common femoral, superficial femoral, popliteal, trifurcation, bypass grafts), presence of visible patent distal vessels, existence of patent collateral vessels, and the occurrence of stenoses involving inflow, collateral, or outflow vessels. Clinical category “viable” (I) correlated with single-segment occlusion, patent collaterals and distal vessels. Patients requiring blood to flow through two contiguous collateral beds to supply distal vessels or with single occlusion combined with stenosis of the adjacent inflow, outflow or collateral vessel correlated with the clinical category “threatened” (II). Occlusions of more than two segments with distal propagation that occluded the distal vascular bed correlated with the clinical category “irreversible” (III). Severity of ischemia and extent of the occlusion correlated with death and amputation rate at 30 days.

In their study of 45 patients with embolic occlusions managed by thrombolysis, Huettl and Soulen^13^ detected no patent calf vessels in 56 % of patients, one calf vessel was patent in 22 %, two in 7 % and three in 16 % of patients before treatment. Those numbers are close to our data. Using thrombolysis and additional endovascular techniques for treatment of non-embolic ALI in 119 patients (72 % grafts, 68 % category I), Kashyap and associates^14^ detected multisegmental thrombosis in 16 % and tibial thrombosis in 22 % of patients. Distal occlusions beyond the reach of the rheolytic catheter limited the treatment in 57 % of 86 patients with ALI or SLI in the study of Kasirajan et al^15^. The need for distal thrombectomy and poor intra-operative runoff were detected among risk factors for amputation and death in 107 patients with ALI undergoing infrainguinal bypass^16^. Also, any patent pedal outflow was a significant predictor for technical success in patients with ALI managed surgically (296 pts) or by endovascular technique (147 pts) in the study of Taha and associates^17^. Failed mechanical recanalization of the diffusely occluded infrapopliteal vascular bed influenced the need for thrombolytic therapy, the rate of major bleeding, treatment outcomes, amputation and mortality rates in patients with ALI and SLI^8^.

The greatest mean lesion lengths were reported in the studies of ALI and SLI patients with target occlusions located in the femoropopliteal bypass (32.1; 28.4 cm)^18,19^. In our series, the mean length of the target occlusion was 22.9 cm and 22. 8 % of 316 patients suffered from occluded femoropopliteal bypass.

The comparison between patients with embolic vs. thrombotic cause of arterial occlusions, we documented higher age, higher frequency of female gender, cardiac disease, arrhythmias, ALI, ALI IIB, occlusions of femoropopliteal segment and/or profunda femoris in subjects with embolism. They also presented higher frequency of additional infrapopliteal occlusion, lower number of patent tibial vessels and lower ABI before treatment. Thus, historical view that embolism produces more severe ischaemia than thrombosis was confirmed.

Thrombosis subgroup presented higher incidence of male gender, smoking history, subacute ischemia, acute ischemia IIA, claudication, tissue defect, femoropopliteal bypass and in-stent occlusions. Furthermore, these patients presented greater mean occlusion length, higher ABI and number of patent calf vessels. Aune and Trippestad^20^ reported in their study of 272 patients managed by embolectomy (192 pts. with embolism) or bypass (80 pts. with thrombosis), a higher mean age, frequency of female gender and cardiac disease, together with shorter life expectancy in patients with embolism. Kuukasjarvi et al. in their study of 509 ALI patients (without previous reconstructions) from National Vascular Registry^21^ detected higher frequency of embolism in patients of age 75 or more years and smoking as a risk factor for acute thrombosis. The authors speculated that the worse prognosis (higher amputation rate, mortality rates and necessity for repeated interventions) of patients with acute thrombotic occlusion was associated with advanced atherosclerosis.

Higher age and frequency of hypertension, arrhythmias, embolism, occlusions of femoropopliteal segment and deep femoral artery were associated with female gender. Smoking history, renal insufficiency, tissue defects, thrombosis as a cause of the occlusion and femoropopliteal bypass occlusion were more frequent in patients of male gender. Additionally, we observed statistical trend toward more frequent claudication in the male group.

Similar results can be found in the literature. Braithwaite and colleagues^22^ managed 84 elderly patients (75-100 years) with ALI by surgery and/or thrombolysis. Percentage of females was 71.4 % in their series and female preponderance was greatest (84.8 %) in the subgroup selected for immediate embolectomy. Female predominance was detected also in other studies: 57 % in subgroup with embolism^18^ and 54 % of 23 268 ALI patients of the National Impatient Sample (NIS)^23^. Kashyap and associates^14^ reported increased risk of limb loss with the female gender in 119 ALI patients managed by thrombolysis. Mortality rate was 6 % and dying patients were women only. Female gender as a risk associated with limb amputation was detected also by Kuoppala M and colleagues^24^. The NATALI study^25^ described female gender as a risk factor for mortality and for all and major complications associated with thrombolytic treatment. On the other hand, Eliason JL et al.^23^ reported decreased risk of amputation in women, in patients with embolectomy, percutaneous transluminal angioplasty and age less than 63 years in NIS patients. The authors did not detect the female preponderance or lower amputation risk in their single institution study (105 patients).

## CONCLUSION

Multisegmental (supratibial plus infrapopliteal) arterial occlusions were documented in 90.6 % of ALI patients and in 78.1 % of SLI patients with threatened lower extremity. The value of ABI related to the number of occluded tibial arteries.

The most frequent locations of supratibial occlusions were femoropopliteal segment (73.1 %) and femoropopliteal bypass (22.8 %) without significant difference in clinical categories. Occlusion locations related to ALI (IIA+IIB) were the aortoiliac segment and/or profunda femoris. Also, more ALI subjects presented additional infrapopliteal occlusion (acute and/or subacute) and lower number of patent tibial arteries and lower ABI.

Anatomically, the important factor related to the ischaemia severity and symptoms duration was the status of infrapopliteal vascular bed. The number of patent or acutely, subacutely and chronically occluded tibial vessels related to individual clinical categories of ALI and SLI and to ABI. This suggests that tibial repefusion can be essential for the salvage of threatened limb and should be always considered.

Patients with the most severe ischaemia and immediately threatened extremity (ALI IIB) presented shortest symptoms duration, higher frequency of rest pain, embolism, diabetes, additional acute infrapopliteal occlusion, lower number of patent tibial vessels and lower ABI in comparison with ALI IIA patients. More frequent diabetes can participate on higher number of occluded tibial vessels in this subgroup.

On the other hand, more patients with thrombosis, smoking history, hyperlipidemia, claudication, higher number of patent calf vessels and higher ABI were detected in ALI IIA subgroup.

In comparison with ALI group, the subjects with SLI presented more frequently the thrombosis as a cause of target occlusion, smoking history, claudication, tissue defects, shorter target occlusion, lower number of occluded tibial vessels and higher ABI. Furthermore, they presented younger age, less embolism and arrhythmias, less aortoiliac segment and deep femoral artery occlusions.

Embolism as a cause of the supratibial arterial occlusion was associated with higher age, female gender, cardiac disease, arrhythmias, ALI, ALI IIB, femoropopliteal segment and deep femoral artery occlusions, infrapopliteal occlusions, lower ABI and number of patent calf vessels. Higher ischaemia severity in subjects with embolism was not associated only with the fact that embolus suddenly occluded healthy supratibial artery but also with higher frequency of additional infrapopliteal occlusion, profunda femoris occlusion and the higher number of occluded tibial vessels. The data confirmed a historical view that the most severe ischemia occurs with the embolism.

Thrombosis was associated with smoking history, SLI, ALI IIA, claudication, tissue defects, longer target lesion, femoropopliteal bypass occlusion, in-stent occlusion, higher ABI and number of patent calf vessels.

Patients of female gender presented higher age, higher frequency of hypertension, arrhythmias, embolism, femoropopliteal and profunda femoris occlusions. Male group manifested a higher frequency of smoking history, renal insufficiency, tissue defect, thrombosis and femoropopliteal bypass occlusion.

Grouping the patients categorized in different clinical categories of ALI and SLI can affect the treatment outcomes.

## Data Availability

available on request

## Acknowledgments

The authors thank Candance McClure (NAMSA, Toledo, OH, USA) and Marian Rybář (MATSTAT, Prague, Czech Republic) for statistical expertise.

## Notes

<u>Declaration of Conflicting Interests:</u> The authors have no potential conflicts of interest with respect to the research, authorship, and/or publication of this article.

<u>Funding</u> The authors recieved no financial support for the research, authorship, and/or publication of this article.

### Competing Interest Statement

The authors have declared no competing interest.

### Clinical Trial

ISRCTN154967770

### Funding Statement

No funding received

### Author Declarations

3rd Medical Faculty, Charles University Centralized Institutional Review Board

